# The race to find a SARS-CoV-2 drug can only be won by a few chosen drugs: a systematic review of registers of clinical trials of drugs aimed at preventing or treating COVID-19

**DOI:** 10.1101/2020.05.05.20091785

**Authors:** Vicente Martinez-Vizcaino, Arthur E. Mesas, Iván Cavero-Redondo, Alicia Saz-Lara, Irene Sequí-Dominguez, Carlos Pascual-Morena, Celia Álvarez-Bueno

**Author notes:** Corresponding author: Dr. Arthur E. Mesas, C/ Santa Teresa Jornet s/n, 16071, Cuenca, Spain.

## Abstract

Considering the massive amount of clinical trial registers aimed to find effective drugs for the prevention and treatment of COVID-19, it is challenging to have a comprehensive view of which drugs are being studied more extensively and when is expected that we will have consistent results regarding their effectiveness. This systematic review included all clinical trials on pharmacological therapy related to COVID-19 and SARS-CoV-2 registered at the International Clinical Trials Registry Platform (WHO-ICTRP) up to April 22, 2020. Clinical trials characteristics (country, design, sample size, main outcomes, expected completion data, type of participants, length of the interventions, main outcomes). How many trials and the accumulated sample size by drug or combination of drugs, and by month in 2020 was depicted. We identified 412 clinical trials registers addressing the effect of pharmacological treatments on COVID-19, predominantly from Asia and Europe (42.2% and 31.1% of clinical trials registers, respectively). The most main outcomes studied were clinical recovery (54.4% of the clinical trials registers, respiratory recovery (28.2%) mortality (27.4%), viral load/negativity (20.4%). During 2020, a huge amount of clinical trials are expected to be completed: 41 trials (60,366 participants) using hydroxychloroquine, 20 trials (1,588 participants) using convalescent’s plasma, 18 trials (6,830 participants) using chloroquine, 12 trials (9,938 participants using lopinavir/ritonavir, 11 trials (1,250 participants) using favipiravir, 10 trials (2,175 participants) using tocilizumab and 6 trials (13,540 participants) using Remdesivir. The distribution of the number of registered clinical trials among the different therapeutic options leads to an excess of sample size for some and a lack for others. Our data allow us to conclude that by the end of June we will have results of almost 20 trials involving 40000 patients for hydroxychloroquine and 5 trials with 4500 patients for remdesivir; however, low statistical power is expected from the 9 clinical trials testing the efficacy of favipiravir or the 5 testing tocilizumab, since they will recruit less than 1000 patients each one.

## INTRODUCTION

The alarming figures of people that have suffered or died by COVID-19 have elicited an impressive pressure on public and private institutions to search drug treatments to prevent and treat the coronavirus. Even the agencies in charge of approving new drugs, or new indications for existing ones, are under extraordinary pressure that could even affect their well-deserved prestige as institutions guided solely and exclusively by scientific rigor in decision-making [1,2]. Investigators and research agencies are determined not to repeat the main mistake made when facing, in 2014, the West African Ebola Epidemic, when, although they produced a plethora of experiments plenty of good intentions, the randomized clinical trials started too late and were not able to recruit enough patients [3].

The spread of the virus appears to have been controlled in Asia and seems to be declining in most European and North American countries; moreover, it is anticipated that summer in the northern hemisphere will significantly reduce the infectivity and virulence of SARS-CoV-2 [4]. However, since no vaccine is expected to be available by then, everything seems to indicate that a resurgence of this pandemic will occur in autumn [5]. As consequence of these hopeless predictions, a desperate race has begun aimed to demonstrate the efficacy of new drugs, and more urgently, to demonstrate that drugs already on the market for other clinical indications such as hydroxychloroquine, remdesivir, azithromycin, etc., are efficacious to manage the SARS-Cov-2 infection [6,7].

In this context, a synthesis effort is lacking to summarize the characteristics of the ongoing clinical trials, the expected dates of the completion of the recruitment, where they are being held, the drugs whose efficacy is being tested and the main outcomes of these trials. Although there are several databases and registries online, there is a concern about when we can expect to have results with a sufficient statistical power to clear up any doubts about the effectiveness of the different therapeutic approaches for the treatment or prevention of the COVID-19 disease.

The objective of this comprehensive systematic review is to gather and synthesize the information included in the clinical trial registers of candidate drugs to prevent and treat COVID-19 according to the pharmacological group and specific drugs name, study design, main outcomes, number and characteristics of participants recruited, and expected completion date. In addition, we graphically represent which drugs are most likely to achieve consistent results over the coming months of 2020.

## METHODS

### Search strategy

Due to the novelty and importance of COVID-19, information regarding unpublished trials was systematically searched through the main clinical trial registries. First, we searched the World Health Organisation - International Clinical Trials Registry Platform (WHO-ICTRP), updated April 22, 2020, as it includes the following primary registries: US National Library of Medicine Registry (Clinicaltrials.gov), Australian New Zealand Clinical Trials Registry (ANZCTR), Brazilian Clinical Trials Registry (ReBec), Chinese Clinical Trial Registry (ChiCTR), Clinical Research Information Service (CRiS) from the Republic of Korea, Clinical Trials Registry - India (CTRI), Cuban Public Registry of Clinical Trials (RPCEC), European Clinical Trials Register (EU-CTR), German Clinical Trials Register (DRKS), Iranian Registry of Clinical Trials (IRCT), International Standard Randomised Controlled Trial Number Registry (ISRCTN) and Japan Primary Registries Network (JPRN). Moreover, the US National Library of Medicine Registry (Clinicaltrials.gov) and ChiCTR, as databases with a higher number of registers, were thoroughly revised themselves in order to avoid leaving out any trial. Clinical trial registries analysing the effect of pharmacological treatments on COVID-19 were eligible. Two authors (AS-L, IS-D) independently screened the trial registries and extracted relevant information. Discrepancies and doubts about relevance of the sources were solved by consensus with two more authors (VM-V, CA-B).

### Clinical trial registries selection

The criteria for the inclusion of clinical trials were as follows: (i) participants— patients with a COVID-19 diagnosis, a close contact with a COVID-19 patient or belonging to a high-risk exposure group such as health careers; (ii) design—randomized control trials (RCTs), nonrandomized experimental studies (non-RCTs) (including at least two-arm pre-post studies), and pilot studies; (iii) type of intervention—studies comparing the effect of pharmacological approaches as treatment or prevention of COVID-19; and (iv) outcomes—mortality, respiratory signs and symptoms’ improvement, oxygen therapy, non-invasive mechanical ventilation [NIMV] and invasive mechanical ventilation [IMV] or extracorporeal membrane oxygenation [ECMO]), clinical recovery (unspecified or scored, clinical, laboratory, imaging recovery or hospital stay-related), viral load/negativity and other outcomes. Furthermore, no restrictions on language, status or year of publication were applied. The criteria for the exclusion of studies were as follows: (i) studies performed in children/adolescents; (ii) non-eligible publication types, such as open-label prospective studies analysing the effect of a pharmacological treatment in COVID-19 or single-arm experimental pre-post studies; and (iii) studies including the effect of stem cells or traditional medicine (Chinese, Iranian, etc.).

### Data Extraction and analysis

The following data were extracted from each register: (1) sample size; (2) location (Africa, Asia, Europe, Latin America, North America, Oceania or ≥2 continents); age of participants (general [≥18 years], only adults [18-65 years] or only elderly [≥65 years]); (3) patients’ status and setting (suspected, confirmed or severe/critical/intensive care unit [ICU]); (4) allocation type (non-RCT or RCT); (5) number of intervention arms (two, three or ≥4 arms); (6) comparison group (placebo/control/usual care, other drugs or both types of comparison groups); (7) phases (I, II, II, IV, I/II, II/III, III/IV); (8) recruitment status (cancelled, recruitment not started, ongoing, completed); (9) intervention length (≥4, 5-8, 9-12, >12 weeks); and (10) estimated completion date.

Clinical trials outcomes (mortality, composite respiratory recovery, signs and symptoms, invasive and non-invasive mechanical ventilation, extracorporeal membrane oxygenation, composite clinical recovery, unspecified or clinical recovery indexes, clinical, laboratory or imaging recovery, hospital stay-related, viral load/negativity and other outcomes) were shown as number and percentage according to the main pharmacological therapies: (1) antivirals (lopinavir/ritonavir, favipiravir, remdesivir); (2) antimalarial (hydroxychloroquine, chloroquine); (3) corticosteroids (methylprednisolone); (4) immunomodulators (tocilizumab, sarilumab); (5) drugs combinations (hydroxychloroquine + azithromycin, hydroxychloroquine + lopinavir/ritonavir); and (6) others (plasma and other drugs or combinations). Additionally, the number of trials expected to be completed and the cumulative sample size expected per month for the year 2020 were shown graphically.

Data extraction was independently performed by two reviewers (IC-R and AEM), and inconsistencies were solved by consensus of two more authors (VM-V, CA-B). The agreement rate between reviewers was reported by calculating kappa statistics.

### Data sharing

The full dataset will be available in online Mendeley Data, a repository of research data that allows to assigns a permanent digital object identifier, un such a way that data of this study can be easily referenced (doi: http://dx.doi.org/10.17632/4rttjzsh4z.1).

## RESULTS

We identified 412 clinical trials registers addressing the effect of pharmacological treatments on COVID-19 (Figure 1), which were conducted in 40 countries, predominantly from Asia and Europe (42.2% and 31.1% of clinical trials registers, respectively), being China (26.5%), USA (13.6%), Iran (10.0%) and Spain (8.7%) the countries with the most registered clinical trials. Clinicals trials sample size ranged from 10 to 100,000 participants, and most clinical trials registers (91.0%) included confirmed COVID-19 patients (13.1% severe/critical clinical status) (Table 1).

**Figure 1.**
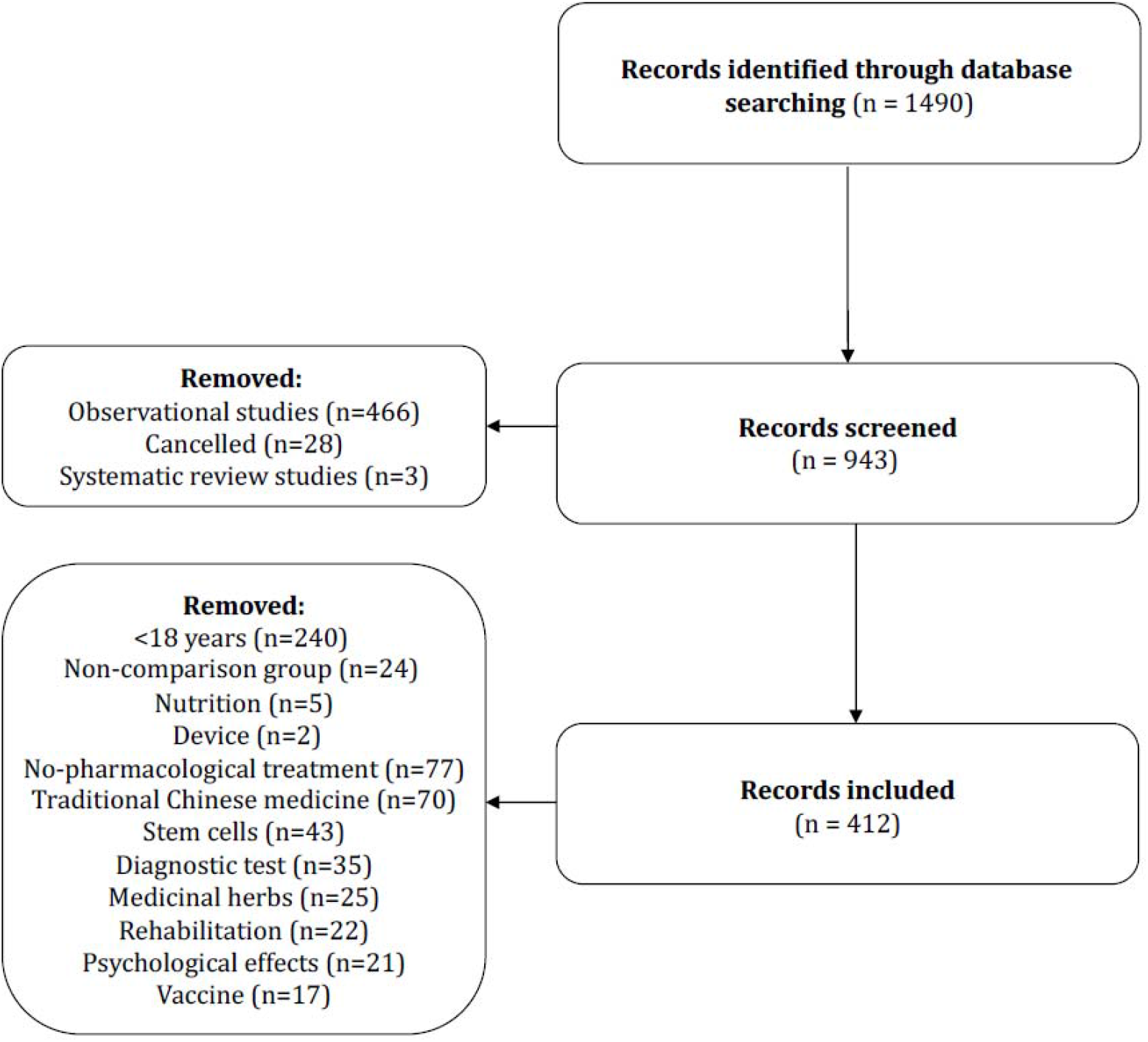
Flow chart

**Table 1.**
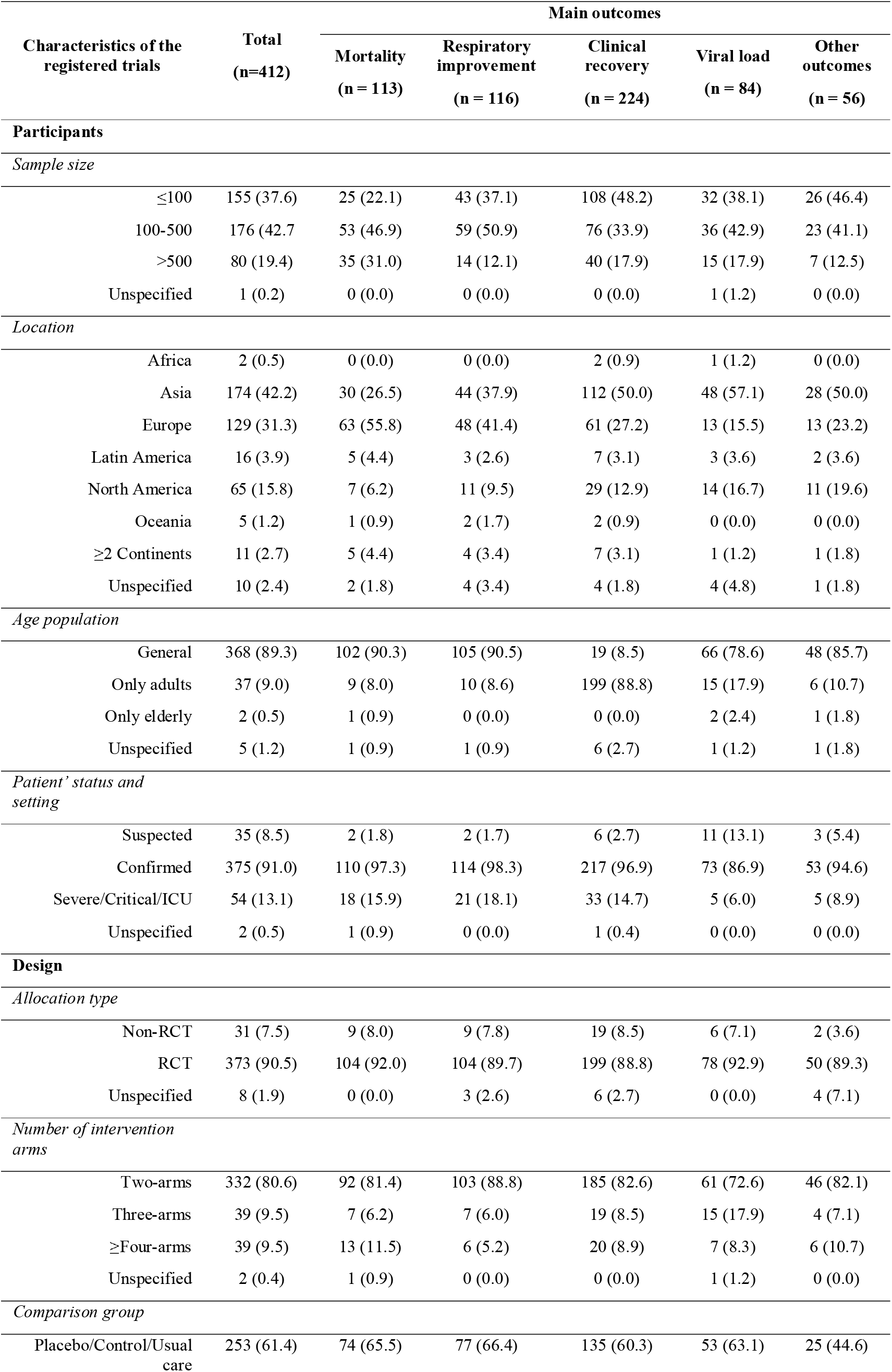
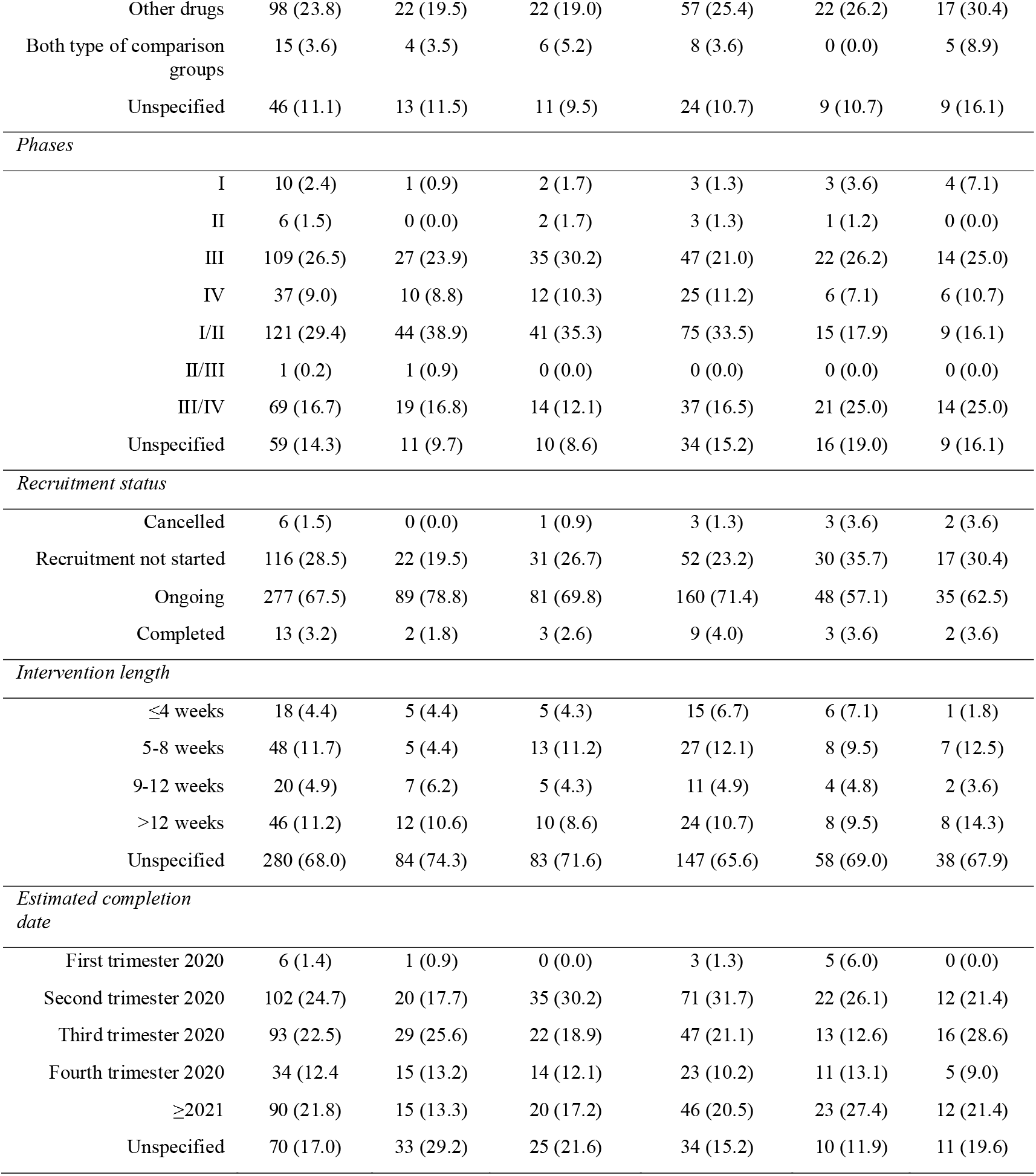
Characteristics of the registered trials aimed to evaluate the effect of pharmacologic strategies to manage the SARS-Cov-19 infection.

| Characteristics of the registered trials | Total<br>(n=412) | Main outcomes |  |  |  |  |
| --- | --- | --- | --- | --- | --- | --- |
|  |  | Mortality<br>(n = 113) | Respiratory improvement<br>(n = 116) | Clinical recovery<br>(n = 224) | Viral load<br>(n = 84) | Other outcomes<br>(n = 56) |
| Participants |  |  |  |  |  |  |
| Sample size |  |  |  |  |  |  |
| ≤100 | 155 (37.6) | 25 (22.1) | 43 (37.1) | 108 (48.2) | 32 (38.1) | 26 (46.4) |
| 100-500 | 176 (42.7) | 53 (46.9) | 59 (50.9) | 76 (33.9) | 36 (42.9) | 23 (41.1) |
| >500 | 80 (19.4) | 35 (31.0) | 14 (12.1) | 40 (17.9) | 15 (17.9) | 7 (12.5) |
| Unspecified | 1 (0.2) | 0 (0.0) | 0 (0.0) | 0 (0.0) | 1 (1.2) | 0 (0.0) |
| Location |  |  |  |  |  |  |
| Africa | 2 (0.5) | 0 (0.0) | 0 (0.0) | 2 (0.9) | 1 (1.2) | 0 (0.0) |
| Asia | 174 (42.2) | 30 (26.5) | 44 (37.9) | 112 (50.0) | 48 (57.1) | 28 (50.0) |
| Europe | 129 (31.3) | 63 (55.8) | 48 (41.4) | 61 (27.2) | 13 (15.5) | 13 (23.2) |
| Latin America | 16 (3.9) | 5 (4.4) | 3 (2.6) | 7 (3.1) | 3 (3.6) | 2 (3.6) |
| North America | 65 (15.8) | 7 (6.2) | 11 (9.5) | 29 (12.9) | 14 (16.7) | 11 (19.6) |
| Oceania | 5 (1.2) | 1 (0.9) | 2 (1.7) | 2 (0.9) | 0 (0.0) | 0 (0.0) |
| ≥2 Continents | 11 (2.7) | 5 (4.4) | 4 (3.4) | 7 (3.1) | 1 (1.2) | 1 (1.8) |
| Unspecified | 10 (2.4) | 2 (1.8) | 4 (3.4) | 4 (1.8) | 4 (4.8) | 1 (1.8) |
| Age population |  |  |  |  |  |  |
| General | 368 (89.3) | 102 (90.3) | 105 (90.5) | 19 (8.5) | 66 (78.6) | 48 (85.7) |
| Only adults | 37 (9.0) | 9 (8.0) | 10 (8.6) | 199 (88.8) | 15 (17.9) | 6 (10.7) |
| Only elderly | 2 (0.5) | 1 (0.9) | 0 (0.0) | 0 (0.0) | 2 (2.4) | 1 (1.8) |
| Unspecified | 5 (1.2) | 1 (0.9) | 1 (0.9) | 6 (2.7) | 1 (1.2) | 1 (1.8) |
| Patient’ status and setting |  |  |  |  |  |  |
| Suspected | 35 (8.5) | 2 (1.8) | 2 (1.7) | 6 (2.7) | 11 (13.1) | 3 (5.4) |
| Confirmed | 375 (91.0) | 110 (97.3) | 114 (98.3) | 217 (96.9) | 73 (86.9) | 53 (94.6) |
| Severe/Critical/ICU | 54 (13.1) | 18 (15.9) | 21 (18.1) | 33 (14.7) | 5 (6.0) | 5 (8.9) |
| Unspecified | 2 (0.5) | 1 (0.9) | 0 (0.0) | 1 (0.4) | 0 (0.0) | 0 (0.0) |
| Design |  |  |  |  |  |  |
| Allocation type |  |  |  |  |  |  |
| Non-RCT | 31 (7.5) | 9 (8.0) | 9 (7.8) | 19 (8.5) | 6 (7.1) | 2 (3.6) |
| RCT | 373 (90.5) | 104 (92.0) | 104 (89.7) | 199 (88.8) | 78 (92.9) | 50 (89.3) |
| Unspecified | 8 (1.9) | 0 (0.0) | 3 (2.6) | 6 (2.7) | 0 (0.0) | 4 (7.1) |
| Number of intervention arms |  |  |  |  |  |  |
| Two-arms | 332 (80.6) | 92 (81.4) | 103 (88.8) | 185 (82.6) | 61 (72.6) | 46 (82.1) |
| Three-arms | 39 (9.5) | 7 (6.2) | 7 (6.0) | 19 (8.5) | 15 (17.9) | 4 (7.1) |
| ≥Four-arms | 39 (9.5) | 13 (11.5) | 6 (5.2) | 20 (8.9) | 7 (8.3) | 6 (10.7) |
| Unspecified | 2 (0.4) | 1 (0.9) | 0 (0.0) | 0 (0.0) | 1 (1.2) | 0 (0.0) |
| Comparison group |  |  |  |  |  |  |
| Placebo/Control/Usual care | 253 (61.4) | 74 (65.5) | 77 (66.4) | 135 (60.3) | 53 (63.1) | 25 (44.6) |
| Other drugs | 98 (23.8) | 22 (19.5) | 22 (19.0) | 57 (25.4) | 22 (26.2) | 17 (30.4) |
| Both type of comparison groups | 15 (3.6) | 4 (3.5) | 6 (5.2) | 8 (3.6) | 0 (0.0) | 5 (8.9) |
| Unspecified | 46 (11.1) | 13 (11.5) | 11 (9.5) | 24 (10.7) | 9 (10.7) | 9 (16.1) |
| *Phases* |  |  |  |  |  |  |
| I | 10 (2.4) | 1 (0.9) | 2 (1.7) | 3 (1.3) | 3 (3.6) | 4 (7.1) |
| II | 6 (1.5) | 0 (0.0) | 2 (1.7) | 3 (1.3) | 1 (1.2) | 0 (0.0) |
| III | 109 (26.5) | 27 (23.9) | 35 (30.2) | 47 (21.0) | 22 (26.2) | 14 (25.0) |
| IV | 37 (9.0) | 10 (8.8) | 12 (10.3) | 25 (11.2) | 6 (7.1) | 6 (10.7) |
| I/II | 121 (29.4) | 44 (38.9) | 41 (35.3) | 75 (33.5) | 15 (17.9) | 9 (16.1) |
| II/III | 1 (0.2) | 1 (0.9) | 0 (0.0) | 0 (0.0) | 0 (0.0) | 0 (0.0) |
| III/IV | 69 (16.7) | 19 (16.8) | 14 (12.1) | 37 (16.5) | 21 (25.0) | 14 (25.0) |
| Unspecified | 59 (14.3) | 11 (9.7) | 10 (8.6) | 34 (15.2) | 16 (19.0) | 9 (16.1) |
| *Recruitment status* |  |  |  |  |  |  |
| Cancelled | 6 (1.5) | 0 (0.0) | 1 (0.9) | 3 (1.3) | 3 (3.6) | 2 (3.6) |
| Recruitment not started | 116 (28.5) | 22 (19.5) | 31 (26.7) | 52 (23.2) | 30 (35.7) | 17 (30.4) |
| Ongoing | 277 (67.5) | 89 (78.8) | 81 (69.8) | 160 (71.4) | 48 (57.1) | 35 (62.5) |
| Completed | 13 (3.2) | 2 (1.8) | 3 (2.6) | 9 (4.0) | 3 (3.6) | 2 (3.6) |
| *Intervention length* |  |  |  |  |  |  |
| ≤4 weeks | 18 (4.4) | 5 (4.4) | 5 (4.3) | 15 (6.7) | 6 (7.1) | 1 (1.8) |
| 5-8 weeks | 48 (11.7) | 5 (4.4) | 13 (11.2) | 27 (12.1) | 8 (9.5) | 7 (12.5) |
| 9-12 weeks | 20 (4.9) | 7 (6.2) | 5 (4.3) | 11 (4.9) | 4 (4.8) | 2 (3.6) |
| >12 weeks | 46 (11.2) | 12 (10.6) | 10 (8.6) | 24 (10.7) | 8 (9.5) | 8 (14.3) |
| Unspecified | 280 (68.0) | 84 (74.3) | 83 (71.6) | 147 (65.6) | 58 (69.0) | 38 (67.9) |
| *Estimated completion date* |  |  |  |  |  |  |
| First trimester 2020 | 6 (1.4) | 1 (0.9) | 0 (0.0) | 3 (1.3) | 5 (6.0) | 0 (0.0) |
| Second trimester 2020 | 102 (24.7) | 20 (17.7) | 35 (30.2) | 71 (31.7) | 22 (26.1) | 12 (21.4) |
| Third trimester 2020 | 93 (22.5) | 29 (25.6) | 22 (18.9) | 47 (21.1) | 13 (12.6) | 16 (28.6) |
| Fourth trimester 2020 | 34 (12.4) | 15 (13.2) | 14 (12.1) | 23 (10.2) | 11 (13.1) | 5 (9.0) |
| ≥2021 | 90 (21.8) | 15 (13.3) | 20 (17.2) | 46 (20.5) | 23 (27.4) | 12 (21.4) |
| Unspecified | 70 (17.0) | 33 (29.2) | 25 (21.6) | 34 (15.2) | 10 (11.9) | 11 (19.6) |

The estimated study completion date ranged from February 2020 to March 2025. Regarding their experimental phase, the registers are distributed in percentages as follows: 2.4%, 1.5%, 26.5% and 29.4% for phases I, II, III and IV respectively. The remaining 14.3% corresponds with those trials including more than an experimental phase or that information is not clearly specified in the register. The allocation of participants to the study groups was done using randomized procedures in the 90.5% of the clinical registers, and 80.6% included two-arms (being placebo/control or usual care the most common comparison group). The intervention length ranged from 1 to 13 weeks. On May 1, only 3.2% of the studies have completed the recruitment (Table 1).

Regarding the COVID-19 outcomes studied, clinical recovery was included in the 54.4%, respiratory recovery in the 28.2%, mortality in the 27.4%, viral load/negativity in the 20.4% and other outcomes in the remaining 13.6%. Furthermore, the most frequent pharmacological treatment under analysis were: hydroxychloroquine (22.8%), chloroquine (6.6%), convalescents plasma (6.6%), lopinavir/ritonavir (5.6%), tocilizumab (4.6%), hydroxychloroquine + azithromycin (4.1%), favipiravir (3.4%), remdesivir (2.9%), methylprednisolone (2.7%), sarilumab (2.4%) and hydroxychloroquine + lopinavir/ritonavir (1.9%) (Table 2). Moreover, during 2020, most of the clinical trials using hydroxychloroquine (41 trials) are expected to be completed, followed by those using plasma (20 trials), chloroquine (18 trials), lopinavir/ritonavir (12 trials), favipiravir (11 trials), and tocilizumab (10 trials) (Figure 2).

**Table 2.**
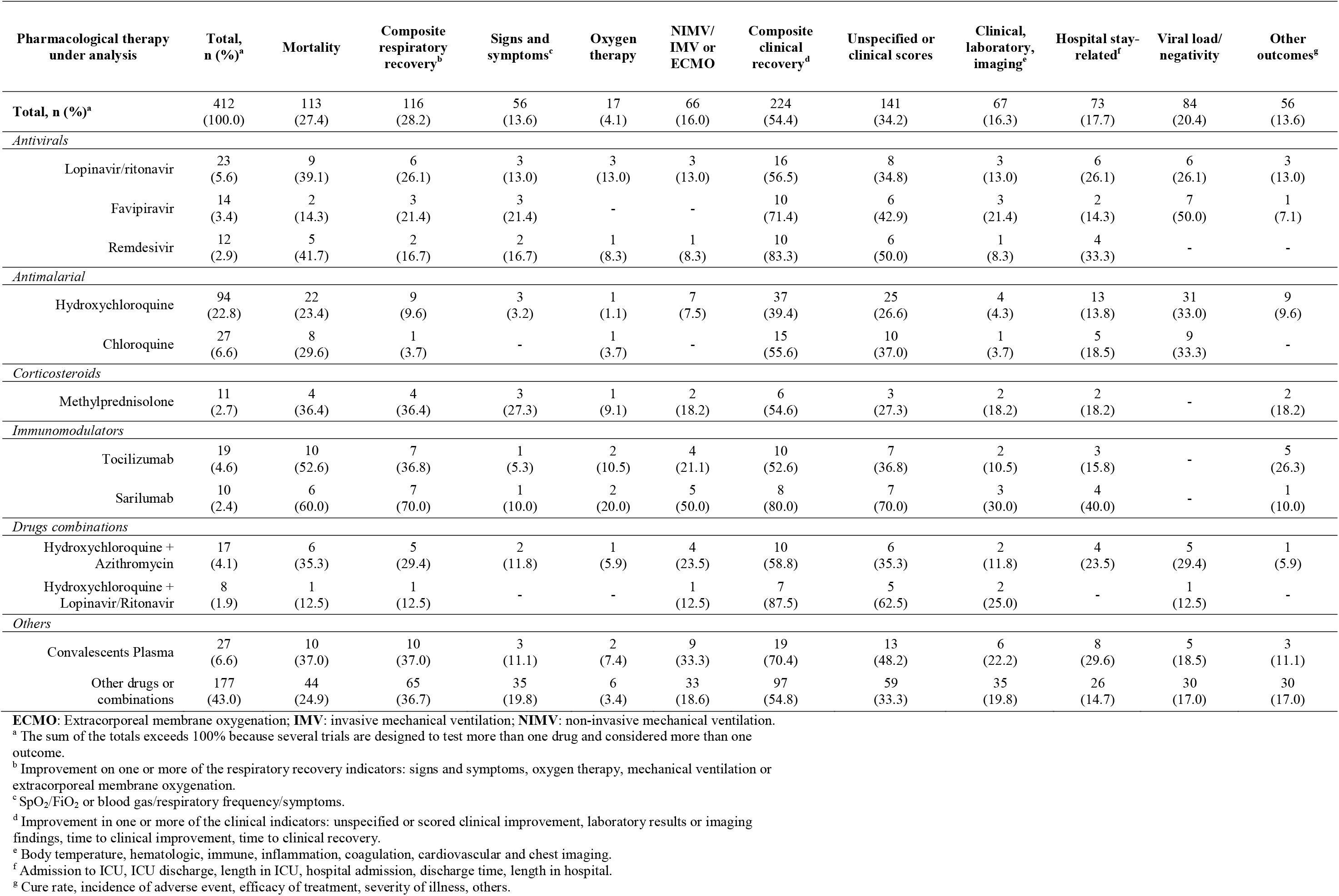
Number (%) of clinical trial registers according to pharmacological therapy and clinical, laboratory or stay-related outcomes.

| Pharmacological therapy under analysis | Total, n (%)<sup>a</sup> | Mortality | Composite respiratory recovery<sup>b</sup> | Signs and symptoms<sup>c</sup> | Oxygen therapy | NIMV/IMV or ECMO | Composite clinical recovery<sup>d</sup> | Unspecified or clinical scores | Clinical, laboratory, imaging<sup>e</sup> | Hospital stay-related<sup>f</sup> | Viral load/negativity | Other outcomes<sup>g</sup> |
| --- | --- | --- | --- | --- | --- | --- | --- | --- | --- | --- | --- | --- |
| **Total, n (%)<sup>a</sup>** | 412 (100.0) | 113 (27.4) | 116 (28.2) | 56 (13.6) | 17 (4.1) | 66 (16.0) | 224 (54.4) | 141 (34.2) | 67 (16.3) | 73 (17.7) | 84 (20.4) | 56 (13.6) |
| *Antivirals* |  |  |  |  |  |  |  |  |  |  |  |  |
| Lopinavir/ritonavir | 23 (5.6) | 9 (39.1) | 6 (26.1) | 3 (13.0) | 3 (13.0) | 3 (13.0) | 16 (56.5) | 8 (34.8) | 3 (13.0) | 6 (26.1) | 6 (26.1) | 3 (13.0) |
| Favipiravir | 14 (3.4) | 2 (14.3) | 3 (21.4) | 3 (21.4) | - | - | 10 (71.4) | 6 (42.9) | 3 (21.4) | 2 (14.3) | 7 (50.0) | 1 (7.1) |
| Remdesivir | 12 (2.9) | 5 (41.7) | 2 (16.7) | 2 (16.7) | 1 (8.3) | 1 (8.3) | 10 (83.3) | 6 (50.0) | 1 (8.3) | 4 (33.3) | - | - |
| *Antimalarial* |  |  |  |  |  |  |  |  |  |  |  |  |
| Hydroxychloroquine | 94 (22.8) | 22 (23.4) | 9 (9.6) | 3 (3.2) | 1 (1.1) | 7 (7.5) | 37 (39.4) | 25 (26.6) | 4 (4.3) | 13 (13.8) | 31 (33.0) | 9 (9.6) |
| Chloroquine | 27 (6.6) | 8 (29.6) | 1 (3.7) | - | 1 (3.7) | - | 15 (55.6) | 10 (37.0) | 1 (3.7) | 5 (18.5) | 9 (33.3) | - |
| *Corticosteroids* |  |  |  |  |  |  |  |  |  |  |  |  |
| Methylprednisolone | 11 (2.7) | 4 (36.4) | 4 (36.4) | 3 (27.3) | 1 (9.1) | 2 (18.2) | 6 (54.6) | 3 (27.3) | 2 (18.2) | 2 (18.2) | - | 2 (18.2) |
| *Immunomodulators* |  |  |  |  |  |  |  |  |  |  |  |  |
| Tocilizumab | 19 (4.6) | 10 (52.6) | 7 (36.8) | 1 (5.3) | 2 (10.5) | 4 (21.1) | 10 (52.6) | 7 (36.8) | 2 (10.5) | 3 (15.8) | - | 5 (26.3) |
| Sarilumab | 10 (2.4) | 6 (60.0) | 7 (70.0) | 1 (10.0) | 2 (20.0) | 5 (50.0) | 8 (80.0) | 7 (70.0) | 3 (30.0) | 4 (40.0) | - | 1 (10.0) |
| *Drugs combinations* |  |  |  |  |  |  |  |  |  |  |  |  |
| Hydroxychloroquine + Azithromycin | 17 (4.1) | 6 (35.3) | 5 (29.4) | 2 (11.8) | 1 (5.9) | 4 (23.5) | 10 (58.8) | 6 (35.3) | 2 (11.8) | 4 (23.5) | 5 (29.4) | 1 (5.9) |
| Hydroxychloroquine + Lopinavir/Ritonavir | 8 (1.9) | 1 (12.5) | 1 (12.5) | - | - | 1 (12.5) | 7 (87.5) | 5 (62.5) | 2 (25.0) | - | 1 (12.5) | - |
| *Others* |  |  |  |  |  |  |  |  |  |  |  |  |
| Convalescents Plasma | 27 (6.6) | 10 (37.0) | 10 (37.0) | 3 (11.1) | 2 (7.4) | 9 (33.3) | 19 (70.4) | 13 (48.2) | 6 (22.2) | 8 (29.6) | 5 (18.5) | 3 (11.1) |
| Other drugs or combinations | 177 (43.0) | 44 (24.9) | 65 (36.7) | 35 (19.8) | 6 (3.4) | 33 (18.6) | 97 (54.8) | 59 (33.3) | 35 (19.8) | 26 (14.7) | 30 (17.0) | 30 (17.0) |

**ECMO:** Extracorporeal membrane oxygenation; **IMV:** invasive mechanical ventilation; **NIMV:** non-invasive mechanical ventilation.
<sup>a</sup> The sum of the totals exceeds 100% because several trials are designed to test more than one drug and considered more than one outcome.
<sup>b</sup> Improvement on one or more of the respiratory recovery indicators: signs and symptoms, oxygen therapy, mechanical ventilation or extracorporeal membrane oxygenation.
<sup>c</sup> SpO<sub>2</sub>/FiO<sub>2</sub> or blood gas/respiratory frequency/symptoms.
<sup>d</sup> Improvement in one or more of the clinical indicators: unspecified or scored clinical improvement, laboratory results or imaging findings, time to clinical improvement, time to clinical recovery.
<sup>e</sup> Body temperature, hematologic, immune, inflammation, coagulation, cardiovascular and chest imaging.
<sup>f</sup> Admission to ICU, ICU discharge, length in ICU, hospital admission, discharge time, length in hospital.
<sup>g</sup> Cure rate, incidence of adverse event, efficacy of treatment, severity of illness, others.

**Figure 2.**
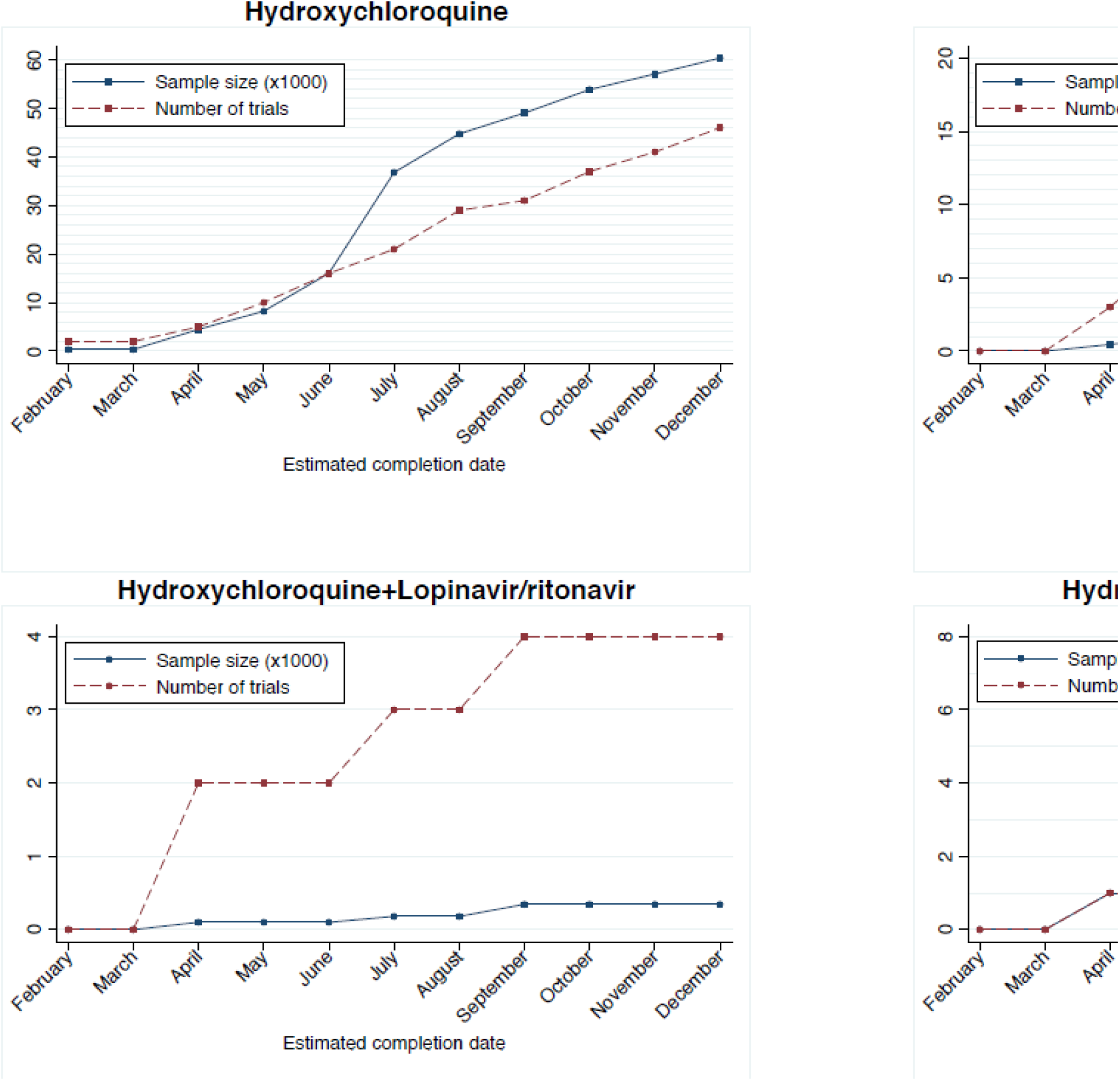
Cumulative sample size and number of clinical trials for antimalarial therapies and combinations by estimated completion date during 2020.

**Figure 3.**
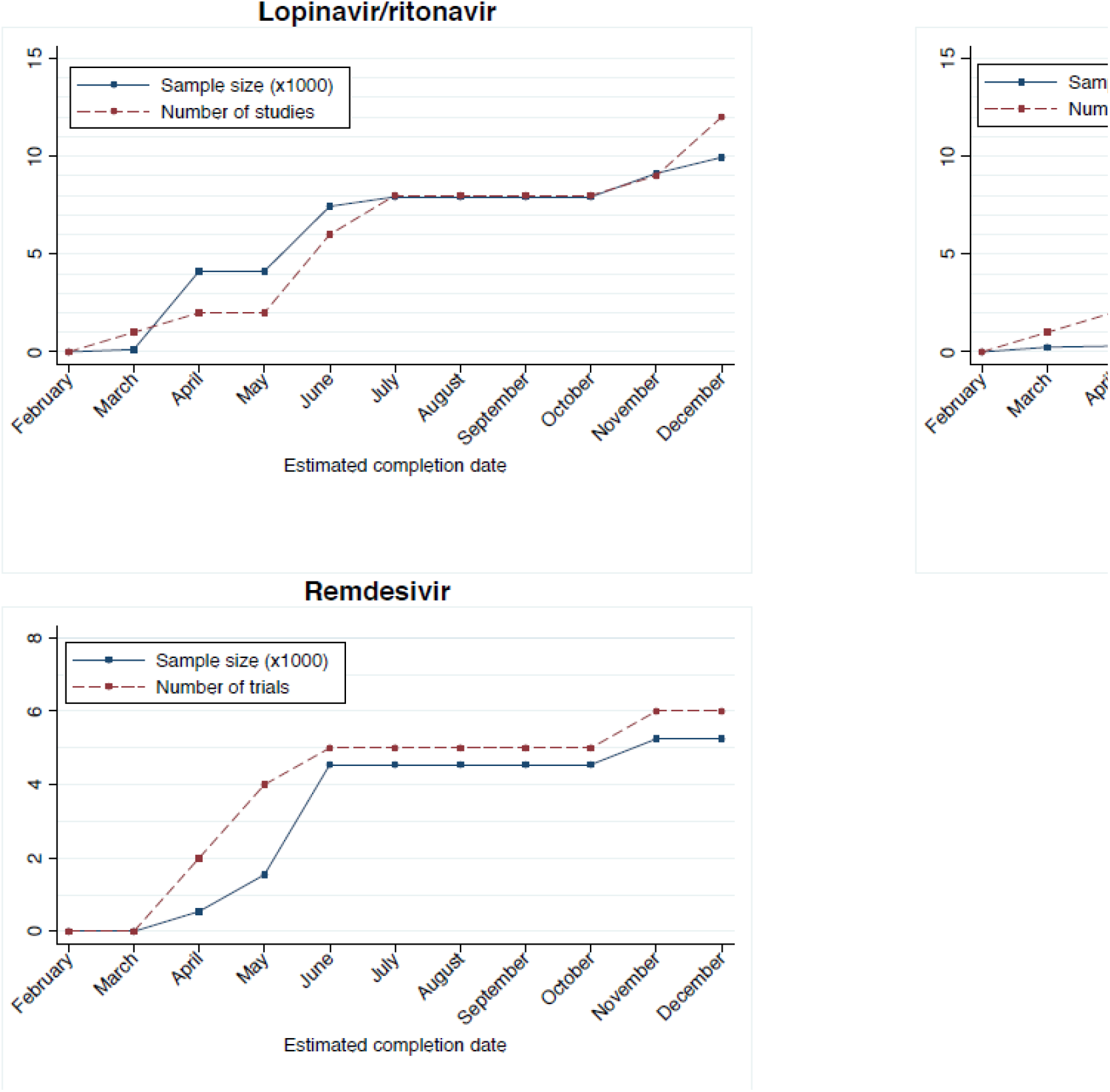
Cumulative sample size and number of clinical trials for antiviral therapies and combinations by estimated completion date during 2020.

**Figure 4.**
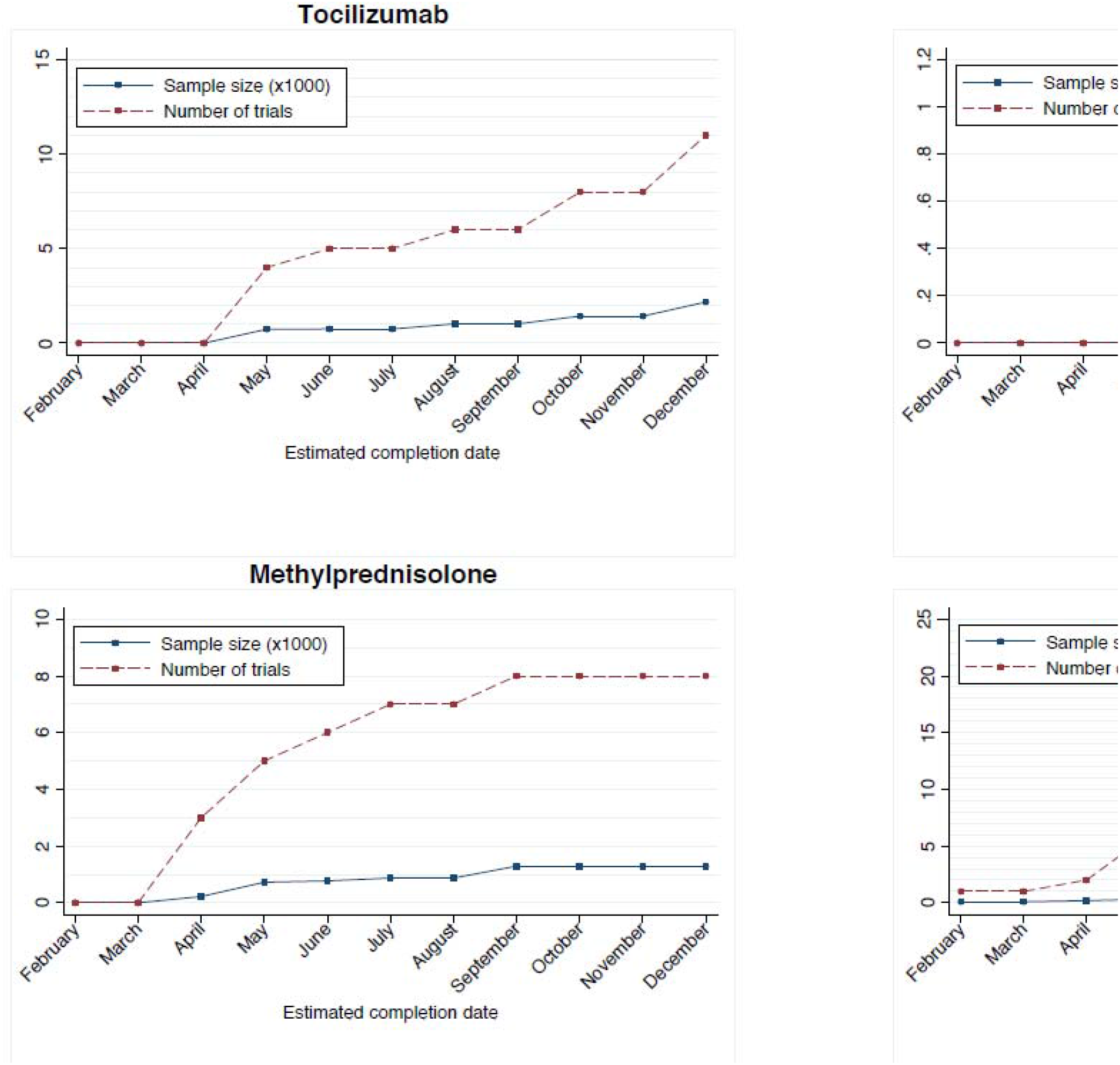
Cumulative sample size and number of clinical trials for immunomodulators, corticosteroid and plasma by estimated completion date during 2020.

Regarding the accumulated sample size by treatment, it is expected that during 2020, the estimated total number of participants to be included in trials with hydroxychloroquine is 60,366 participants, with lopinavir/ritonavir 9,938, with chloroquine 6,830, with remdesivir 5,245, with hydroxychloroquine + azithromycin 3,401, with tocilizumab 2,175, with convalescents plasma 1,588, with methylprednisolone 1,292, with favipiravir 1,250, with hydroxychloroquine + lopinavir/ritonavir 345, and only 60 with sarilumab (Figure 2). Additionally, the number of participants will significantly increase beyond 2021 for some treatments, such as hydroxychloroquine (93,630 participants), lopinavir/ritonavir (20,198 participants), remdesivir (13,540 participants), chloroquine (12,160 participants), sarilumab (2,140 participants) and hydroxychloroquine + azithromycin (1,376 participants). It should be noted that these figures may be underestimated because many registers did not indicate the estimated completion date, which would increase the sample size for hydroxychloroquine + lopinavir/ritonavir in 3,915 participants, for hydroxychloroquine+azithromycin in 2,252 participants, for sarilumab in 2,206, and for favipiravir in 1,107.

## DISCUSSION

The world is currently facing a SARS-CoV-2 pandemic for which there are not effective therapeutic resources yet available. So much so that most hospitalized patients with COVID-19 diagnosis have had to receive off-label or compassionate use therapies, because there were not drugs approved for this disease [8,9]. Because a resurgence of this pandemic seems likely to happen in the autumn [10], it is urgent to test the effectiveness of new drugs, or repurposed ones, for preventing and treating the COVID-19 disease [11,12]. In this context, this study provides an overview of the characteristics of the worldwide registered clinical trials until April 22, 2020.

The main characteristics of the registered trials are the following: more than half are aimed at recovery of clinical or respiratory parameters, are conducted in in China, Europe and USA, participants are mostly hospitalized patients with confirmed COVID-19, have small/moderate sample size, up to 68.0% of trials have not specified the duration of the intervention in their registers, and up to 34.2% are going to end after the fall. Moreover, most trials are testing the efficacy of antimalarial drugs, particularly hydroxychloroquine (alone or in combination with azithromycin or lopinavir/ritonavir) and chloroquine, followed by the antiviral drugs lopinavir/ritonavir, favipiravir and remdesivir. Indeed, substantial interest has been placed in demonstrating the effectiveness of these specific drugs or drugs combinations [13-15].

The challenge for which most of these clinical trials have been planned is to prevent or mitigate the devastating impact of the SARS-CoV-2 infection on the world’s health while an effective vaccine emerges. And the obvious question is whether all this enormous deployment of research resources will not be a futile and disheartening effort. If we consider the cumulative sample size, the number of studies and the estimated date for completing the recruitment of participants for each of the treatment options (Figure 2), the current results seem to indicate that this risk is real. Thus, we can observe that by the end of June we will most likely to know whether hydroxychloroquine, chloroquine, remdesivir, or the combinations of hydroxychloroquine with azithromycin or lopinavir/ritonavir are effective to manage COVID-19 cases, because the ongoing trials will have accumulated, for each of these substances, results from more than 3000 patients, and in some cases as for hydroxychloroquine more than 15000 patients. Therefore, if the data of the registers are accurately completed, as scheduled many of the trials to assess the effectiveness of these therapeutic options planning to recruit patients after June could be cancelled or readapted.

In this context, it is worth highlighting two major recent research initiatives which, in parallel, aim to produce the same results. The Solidarity Trial [16], sponsored by the WHO, which will involve more than hundred countries, and almost simultaneously, the European trial Discovery [17], financed mainly through European institutions and governments. Both mega-trials are aimed to evaluate the efficacy of Remdesivir, Chloroquine or hydroxychloroquine, Lopinavir/Ritonavir and Lopinavir/Ritonavir plus Interferon beta-1 in patients hospitalized with COVID-19. Both are pragmatic trials and, because of their adaptive design, will allow incorporating new therapeutic options during their development. Although through meta-analysis techniques the results of these trials can be synthesized as if they were a unique trial, it seems thoughtful that it would have been more efficient to coordinate these research efforts, gaining statistical power and, perhaps, to include some promising therapeutic alternatives not as extensively examined as favipiravir or tocilizumab.

Finally, the analysis of the data from the 412 clinical trials registered up to April 22, 2020 it can be summarized as follows: i) most of the trials are aimed at repurpose drugs or combinations of these drugs; ii) for all these therapeutic options the cumulative sample size seems excessive, albeit at expense of some small clinical trials with questionable quality in their registers; iii) most likely in June we will have consistent evidence about the effectiveness of the most promising therapeutic options (hydroxychloroquine, remdesivir, Lopinavir/Ritonavir), and with less certainty we will also know if favipiravir, tocilizumab, methylprednisolone, convalescent’s plasma are useful to improve the clinical course of the COVID-19 disease; iv) it is unlikely that by the autumn resurgence of the illness we will have the results of the trials that are testing the efficacy of new drugs against the SARS-CoV-2 infection.

Among the limitations of this study it should be acknowledged that: i) the quality of the registers has not been evaluated due to the lack of a reliable tool for this purpose; ii) although the WHO-ICTRP is the main database of clinical trial registers and collects most national and international databases, some trials registered in non-WHO-ICTRP databases may not have been included; iii) the information collected from each clinical trial register could be heterogeneous, because each database encodes the information differently; iv) because of the lack of quality in the description of the sources of funding in many registers, we have not examined the potential relationship between therapeutic options and public or private sponsors of the trials; and v) the figures provided relating number of trials with sample size estimated to be available during 2020 may be underestimated, because many registers do not indicate the estimated end date of the trial.

In summary, regardless of whether it is financed by public or private resources, and especially in this pandemic crisis, research is a public good, and therefore has to be based on its scientific and social value, in order to produce the scientific evidence needed by the population, policy makers and, above all, clinicians [18]. For this reason, it is not justified that predictably by June more than 20 clinical trials enrolling tens of thousands of patients will finish their recruitment processes testing almost identical hypotheses, thus neglecting other promising therapeutic hypotheses. Probably, as has been recently suggested [19], this pandemic context it is the right time to demand from academic biomedical centres and supranational public health institutions led the coordination of the research initiatives, otherwise much of the research on the management of SARS-CoV-2 will be a waste of research resources [20]. Finally, in view of the low level of precision with which many of the database items have been recorded, a quality assessment tool for clinical trial records becomes necessary, since accuracy of these records would predict the risk of bias of the trial’s results.

## Data Availability

All data come from public domain International Clinical Trials Registry Platform (WHO-ICTRP).

